# Community -Based Intervention to reach the last mile of childhood vaccination coverage in Rwanda: Case study of Burera district

**DOI:** 10.1101/2025.05.23.25328222

**Authors:** Michee Nshimayesu, Barnabas Tobi Alayande, Sage M. C. Ishimwe, Aimable Uwimana, Charif Niyiragira, Ernest Tambo

## Abstract

**Background:** Vaccination remains a cornerstone of global child health, offering one of the most cost-effective interventions for reducing childhood morbidity and mortality caused by vaccine-preventable diseases (VPDs). Despite this, global vaccination coverage has recently declined, particularly in low- and middle-income countries. In Rwanda, although the national childhood immunization rate has reached 95%, gaps remain in reaching the last mile. Community Health Workers (CHWs) have played an instrumental role in promoting and delivering immunization services across hard-to-reach communities. Yet, little is known about the specific contributions of CHWs to these gains, particularly in rural settings such as Burera District, Northern Rwanda. The study aimed to (1) investigate the factors influencing CHWs’ intervention in promoting childhood vaccination, (2) examine the impact of CHWs on vaccination coverage among children under five, and (3) explore the perspectives of caregivers on CHWs’ intervention in childhood vaccination coverage.

**Methods:** A mixed-methods approach was adopted, combining quantitative surveys with 345 CHWs and qualitative key informant interviews (KIIs) with selected caregivers of children under five benefiting from CHWs vaccination intervention. Quantitative data were collected through structured questionnaires and analyzed using descriptive statistics, and multivariable logistic regression in STATA 18. Thematic analysis was used to analyze qualitative data obtained from five KIIs across Burera’s five major health center catchment areas.

**Results:** CHWs in Burera were predominantly female (67.5%), married (98.8%), with primary-level education (86.1%), and most engaged in farming (98.6%). A large proportion (94.5%) had received training on vaccination services, and 98.3% regarded under-five vaccination as “very important.” Over 50% reported facilitating community vaccination events twice per month. CHWs were actively involved in community mobilization and advocacy, with 93.9% participating in campaign planning. CHW activities included referring children to health facilities (58.0%) and conducting home-based vaccination outreach (42.0%). From caregivers’ perspectives, CHWs played critical roles in health education, timely reminders for appointments, and home-based vaccination services. Thematic analysis of interviews revealed strong trust in CHWs, who were seen as vital liaisons between the community and the formal health system. Barriers noted included logistical challenges and the need for more consistent training and communication support for CHWs. Over 55% of CHWs recommended more structured community awareness campaigns to further enhance immunization uptake.

**Conclusion:** CHWs are central to Rwanda’s success in achieving high childhood vaccination coverage, especially in rural areas like Burera District. Their interventions in education, mobilization, and service delivery have significantly contributed to bridging gaps in access and awareness. However, sustaining and enhancing these gains requires continued investment in CHW training, supervision, and logistical support. Findings from this study provide critical insights for policymakers and health planners to strengthen community-based vaccination strategies and reach the last mile by 2030 in alignment with Sustainable Development Goal (SDG) 3.b.

## BACKGROUND

Vaccination is one of the most effective and cost-efficient public health interventions for reducing child morbidity and mortality, particularly from vaccine-preventable diseases (VPDs)^1^. However, vaccination coverage remains a concern globally. In 2023, global immunization coverage showed mixed progress: 14.5 million children remained completely unvaccinated (zero-dose), diphtheria, tetanus, and pertussis (DTP3) and first-dose measles coverage plateaued at 84% and 83% respectively (below pre-pandemic levels), HPV first-dose coverage in girls improved from 20% to 27%, and yellow fever vaccination in at-risk countries lagged at 50%, far short of the recommended 80% threshold^2^. In 2019, an estimated 5.3 million children under the age of five died, nearly half (49.2%) of them from infectious causes, primarily preterm birth complications, lower respiratory infections, intrapartum-related events, and diarrhoea. Notably, 21.7% of these deaths were caused by vaccine-preventable diseases such as measles, meningitis, and respiratory infections, while many more could have been prevented with low-cost interventions. Since 2000, under-5 mortality has declined significantly, owing primarily to reductions in deaths from these preventable and treatable conditions^3^.

Community Health Workers (CHWs) play a vital role in improving child health outcomes, especially in underserved communities. Their involvement in delivering curative care for malaria, pneumonia, and diarrhea has been instrumental in reducing childhood illness and mortality. In Sub-Saharan Africa, CHW-led interventions which decrease in child mortality through direct care, health education, and underscoring their critical contribution to closing healthcare gaps. CHWs identified and referred 42 probable measles cases for immunization, while in Kenya, CHW programs increased infant vaccination rates from 88.7% to 98.8%^4, 5, 6^.

Rwanda implemented its CHW program in 1995 to enhance healthcare access and address medical professional shortages^7^. Since 2015, the number of CHWs has increased to four per village, with two-thirds being women, significantly contributing to health interventions, including childhood vaccinations^8^. CHWs have been instrumental in improving vaccination rates, contributing to Rwanda’s 95% childhood immunization coverage in alignment with Sustainable Development Goal (SDG) 3.b ^9, 10, 11^.

Despite Rwanda’s success, little is known about the specific contributions of CHWs to national vaccination achievements. We sought 1) to investigate the factors influencing the intervention of CHWs in promoting childhood vaccination, 2) to examine the effects of CHWs on the childhood vaccination coverage, and 3) to explore the perspectives of mothers/caregivers on CHWs’ intervention in childhood vaccination coverage in Burera district, Rwanda.

## METHODS AND MATERIALS

### Study design, setting and study population

This research employed a mixed-methods approach, combining quantitative and qualitative data collection and analysis techniques. This approach enabled a comprehensive understanding of the contribution of CHWs’ intervention in the under-5 children’s vaccine program in Burera district communities. This study was conducted in Burera District which is divided into 17 sectors with a total of 69 cells and 571 villages distributed in all sectors, in the Northern Province of Rwanda. Burera district occupies 645 km^2^ and accounts for 397,754 habitants (199,706 males and 176,187 females), which represents 18% of the total population of the Northern province^12^.

This study focused on CHWs and under-5 mothers in the Burera District of Rwanda’s Northern Province. Burera district has a total of 2284 CHWs. Each village, which has 100-250 households, has four CHWs, one female “Agent de Sante Maternelle (ASM)” and a male-female pair known as “Binômes.” ASMs provide maternal and newborn healthcare through home visits and follow-up, whereas “Binômes” provide a broader range of services, including childhood illness diagnosis and treatment, antimalaria services for people of all ages, malnutrition screening, and contraceptive provision.^11,13^. Additional to the quantitative feedback from CHWs, the study focused on the mothers or caregivers’ perceptions who have children benefiting from both vaccination services and CHWs intervention to collect the qualitative insights of services beneficiaries in Burera community.

### Inclusion and Exclusion Criteria

The inclusion criteria for quantitative participants, included being a CHW living and working in Burera district, or having been a CHW in the same region for at least two years, and voluntary consenting to participate in the study. CHWs in charge of health services other than the vaccination excluded in the study, and the CHWs who are not active in the services during the study period were excluded For qualitative participants, we included mothers or primary caregivers of children under-five years who had, lived in the specific village for at least 2 years Mothers who did not use in community based-vaccination services or CHW services, and those unwilling to participate in the study were excluded a.

### Sample size calculation and Sampling technique

To calculate the quantitative participants from 2284 CHWS living and working in Burera, we calculated the sample size using a 95% confidence level with a 5% margin of error. Burera now has 5% of Rwanda’s CHWs (2,284/45,516 total). Using Slovin’s formula, we arrived at a minimum sample size of 340 CHWs^14^. A sampling frame technique used to select participants out of 2284 CHWs present in Burera district. Lists of CHWs were obtained from health centers, and a simple random sampling technique was applied every fourth name on the list was selected in each health center. Additionally, we targeted the health centers with the highest number of CHWs to ensure broader representation, inviting equal numbers of participants from each selected center. For the qualitative component, five caregivers were selected one from each of the five health centers in Burera district. Using a list provided by each health center, we randomly selected every fourth individual who met the eligibility criteria within the respective catchment area. To ensure participants met the study selection criteria, a purposive sampling approach was also applied to identify mothers or caregivers of under-five children who were actively participating in community-based vaccination services. The lists obtained from health centers served as the basis for randomly selecting eligible participants.

### Data collection

A data collection process took place between May 06^th^ 2024 to June 16^th^, 2024. The trained data collector was hired to facilitate the data collection process. Initially, participants’ information, including contact details, was extracted from health center records. The data collector then contacted the participants to obtain a written consent before they decided whether or not to participate. For quantitative data collection, a structured questionnaire was developed and administered to CHWs. This questionnaire aimed to gather information on the factors influencing their interventions, as well as their roles and impact on the vaccination program. It also collected demographic data about the CHWs. The quantitative data collection involved face-to-face administration of close-ended questionnaires, which consist of a series of questions designed to elicit specific responses from participants^15^. In addition, KIIs were conducted with caregivers to explore their experiences and perceptions related to the childhood vaccination program and the interventions of CHWs. The KIIs were audio-recorded and subsequently transcribed for thematic analysis.

### Study Variables and measures

The study examined the perceived effectiveness of CHWs’ interventions on childhood vaccination coverage, considering various independent variables. These included socio-demographic factors, infrastructure and logistics, training, knowledge, community awareness and perception, community mobilization, services available at the CHW level, availability of health information and communication for under-5 mothers, CHWs’ availability for home visits, and equitable service delivery for under-5 children and their mothers. The study measures focused on three key aspects of CHW’s interventions in childhood vaccination services: availability, accessibility, and quality. These aspects were examined through two main approaches. The first approach assessed community awareness and perceptions of CHWs’ interventions. The second approach evaluated changes in the likelihood of under-5 children being fully vaccinated before and after CHWs’ interventions. Accessibility was analyzed based on location, communication, appointment availability, service costs, frequency of access, access to medical records, and age-related access. Quality was assessed through confidentiality and privacy during visits, service provision, accessibility, environmental factors, adherence to written guidelines, CHW characteristics and competencies, and the involvement of under-5 mothers and caregivers.

### Data management

All participants were required to sign a consent form approved by the UGHE IRB Committee before the start and those signed the consent form assigned a unique study ID code to be used throughout the study period. During data collection, the information was cross-checked and further verified by the principal investigator to ensure data quality and integrity. Quantitative data was collected using REDCap (Research Electronic Data Capture) and subsequently transferred to STATA (Stata Corp Inc.) version 18 for analysis (Ref). Key Informant Interviews (KIIs) were audio-recorded and subjected to thematic analysis to identify common themes and patterns related to the interventions of community health workers (CHWs) on vaccination coverage. All data files associated with the study were securely stored on the researchers’ computers, protected by strong passwords.

### Data analysis

Quantitative data were checked for completeness, edited, coded, and exported for analysis into STATA (Stata corp inc.) version 18. Descriptive statistics were performed for prevalence estimation and participant demographic characteristics after cleaning the data for inconsistencies. To determine the relationship between different variables, the Chi-square test, and the t-test were used. Also, multivariable analyses used to investigate the relationship between under-5 vaccination coverage and CHW intervention. A statistically significant p-value of 0.05 was considered during the analysis. For qualitative data (KIIs), thematic analysis was employed to identify common themes and patterns in the data. Transcripts were coded accordingly, and codes were then grouped into themes. The findings are presented using narrative descriptions, illustrative quotes, and thematic matrices.

## RESULTS

### Socio-demographic characteristics of the community health workers

The highest percentage (38.8%) of the community health workers (CHWs) were aged between 40 and 49 years whereas only a small proportion (7.5%) were aged less than 30 years. Females constituted the majority (67.5%) and about half were identified as Catholic (48.4%). Al most all (98.8%) were married and 86.1% of them attained primary level of education. A large percentage (98.6%) were engaged in farming (Table 1).

**Table 1.**
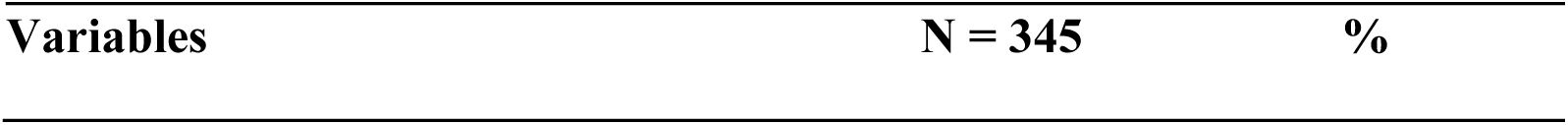
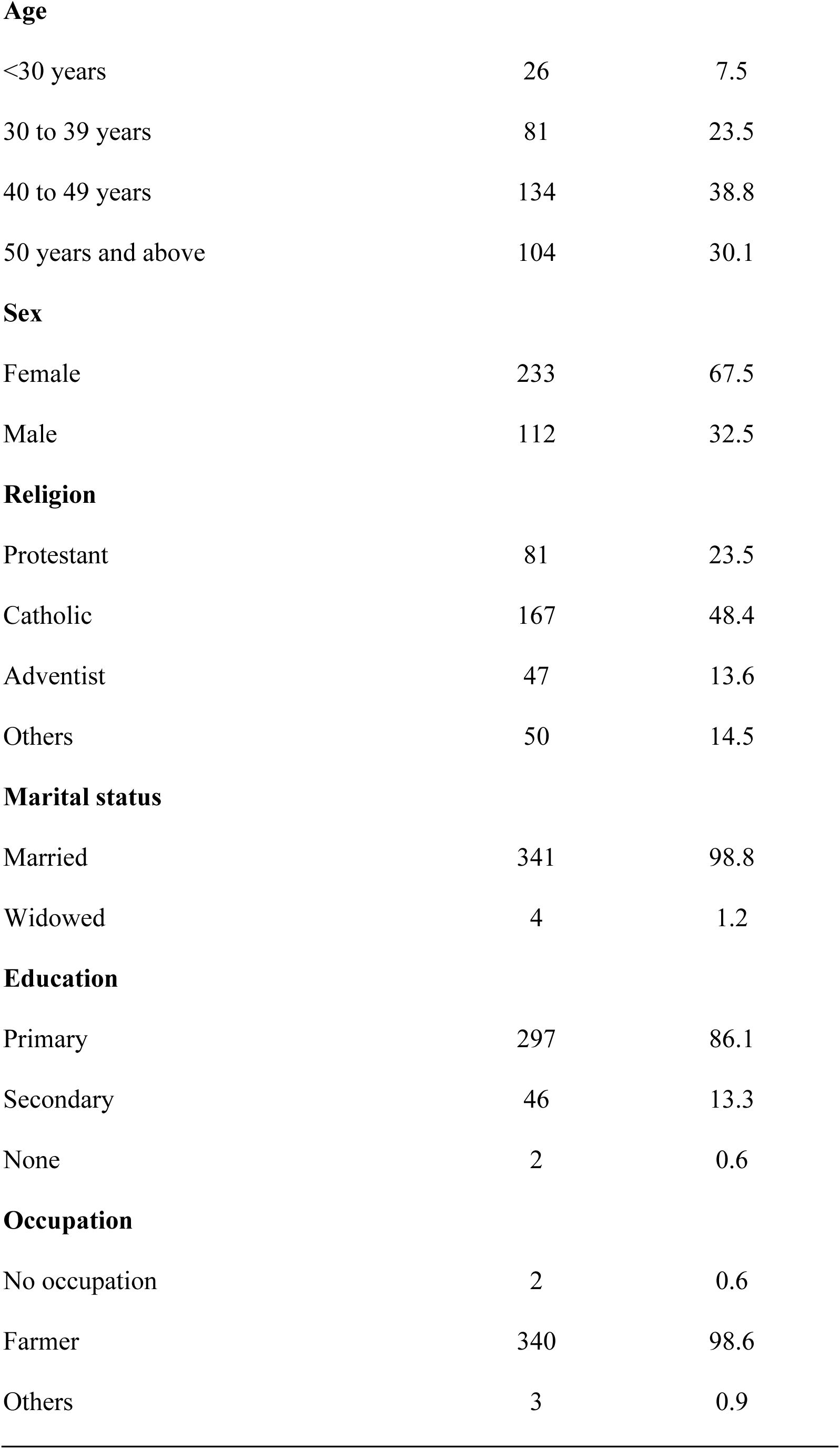
Socio-demographic characteristics of community health workers.

| Variables | N = 345 | % |
| --- | --- | --- |
| **Age** |  |  |
| <30 years | 26 | 7.5 |
| 30 to 39 years | 81 | 23.5 |
| 40 to 49 years | 134 | 38.8 |
| 50 years and above | 104 | 30.1 |
| **Sex** |  |  |
| Female | 233 | 67.5 |
| Male | 112 | 32.5 |
| **Religion** |  |  |
| Protestant | 81 | 23.5 |
| Catholic | 167 | 48.4 |
| Adventist | 47 | 13.6 |
| Others | 50 | 14.5 |
| **Marital status** |  |  |
| Married | 341 | 98.8 |
| Widowed | 4 | 1.2 |
| **Education** |  |  |
| Primary | 297 | 86.1 |
| Secondary | 46 | 13.3 |
| None | 2 | 0.6 |
| **Occupation** |  |  |
| No occupation | 2 | 0.6 |
| Farmer | 340 | 98.6 |
| Others | 3 | 0.9 |

### Knowledge and training on childhood vaccination program

Most of the CHWs (50.4%) reported that vaccination services are offered twice a month. The majority (94.5%) of CHWs had received education or training on vaccination services with 64.1% having participated in training not more than twice. Significant percentage (98.3%) indicated that vaccination under 5 years is very important and 94.2% reported that missing vaccination could cause diseases (Table 2).

> This was also supported in the qualitative data as one pregnant woman expressed “*Community health workers explain it well each time we come for vaccination, starting from when you get tested for pregnancy, they explain how you have to get your child vaccinated on time*” (KII-3).
>
> Another woman also endorsed this as “*They inform and educate us on how to vaccinate our children, starting from the beginning of pregnancy. As soon as you find out you are pregnant, they tell you that you will give birth at a health facility and follow up with the first vaccination. They ensure vaccinations at one and a half months, two and a half months, and continue at nine and fifteen months*” (KII-4).

**Table 2.** Knowledge and training on childhood vaccination program.

| Variables | N = 345 | % |
| --- | --- | --- |
| **Frequency of offering vaccination services in one month** |  |  |
| 0 time | 15 | 4.3 |
| 1 time | 69 | 20.0 |
| 2 times | 174 | 50.4 |
| 3 times | 44 | 12.8 |
| 4 times | 27 | 7.8 |
| 5+ times | 16 | 4.6 |
| **Ever received any education or training about vaccination services** |  |  |
| Yes | 326 | 94.5 |
| No | 19 | 5.5 |
| **Frequency of training received about vaccination services in year (n = 326)** |  |  |
| 1-2 times | 209 | 64.1 |
| 3 - 4 times | 92 | 28.2 |
| 5 times and above | 25 | 7.7 |
| **How important is the under-5 vaccination program in the community** |  |  |
| Not important | 1 | 0.3 |
| Important | 5 | 1.4 |
| Very important | 339 | 98.3 |
| **Main effects of missing vaccine for under-5** |  |  |
| Cause disability | 23 | 6.7 |
| Cause death | 1 | 0.3 |
| Cause of disease | 325 | 94.2 |
| Don't know | 1 | 0.3 |

### Community mobilization and participation towards childhood vaccination program

As indicated in Table 3, the majority of CHWs (93.9%) were fully engaged in conducting vaccination program campaign organized by community leaders during planning phase (Table 3). This is reinforced by the qualitative data where one pregnant woman indicated:

> “*They encourage us to be there, they pay us a visit to let us know that we will be meeting on the vaccination site and do what is required from us* (KII-3).
>
> Another woman also stated, “*Community health workers manage it efficiently. When it comes to pills or drops, they are available at the local service centers. They maintain a list of children and ensure each child receives the appropriate vaccines according to their area (KII-2)*.

**Table 3.**
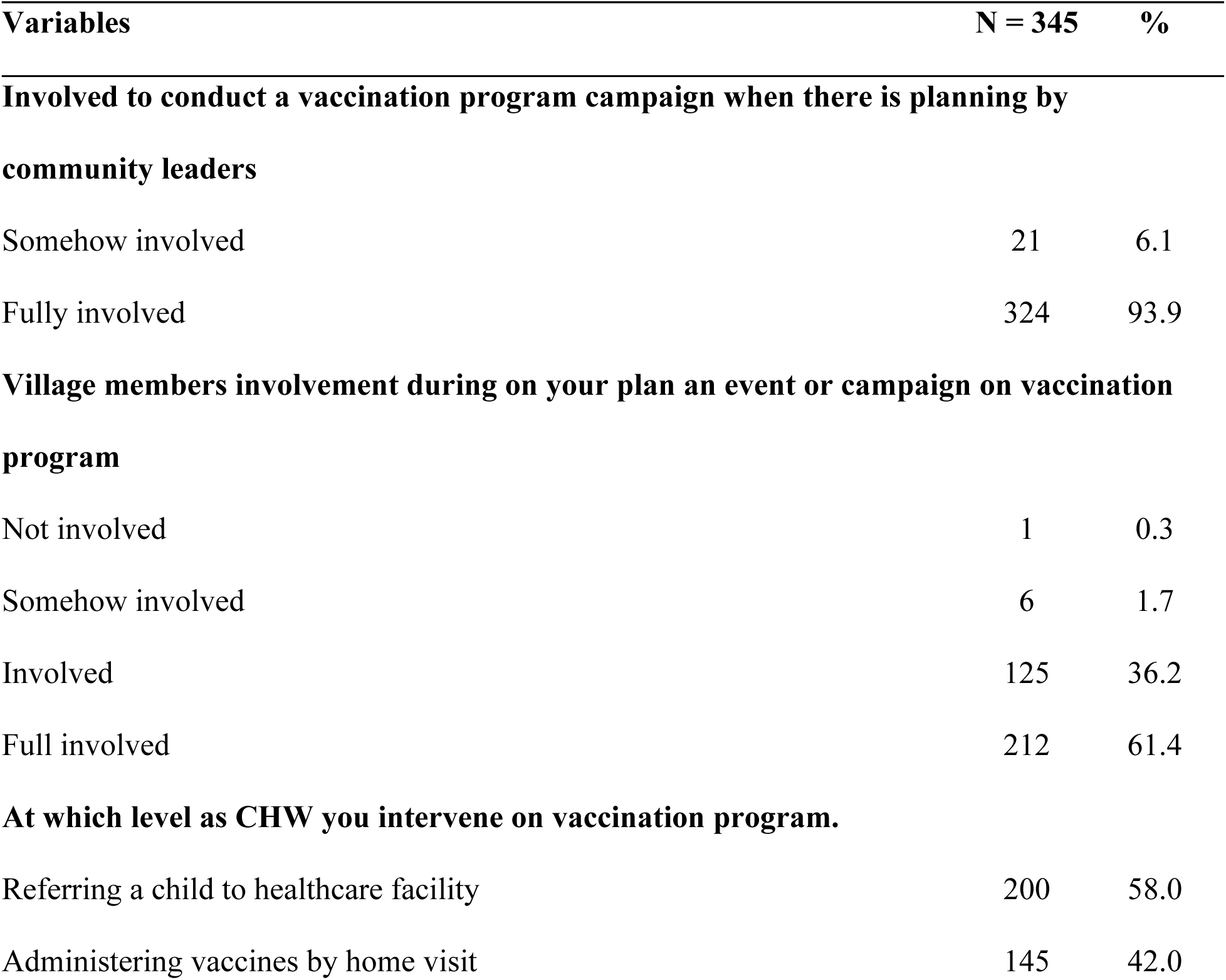
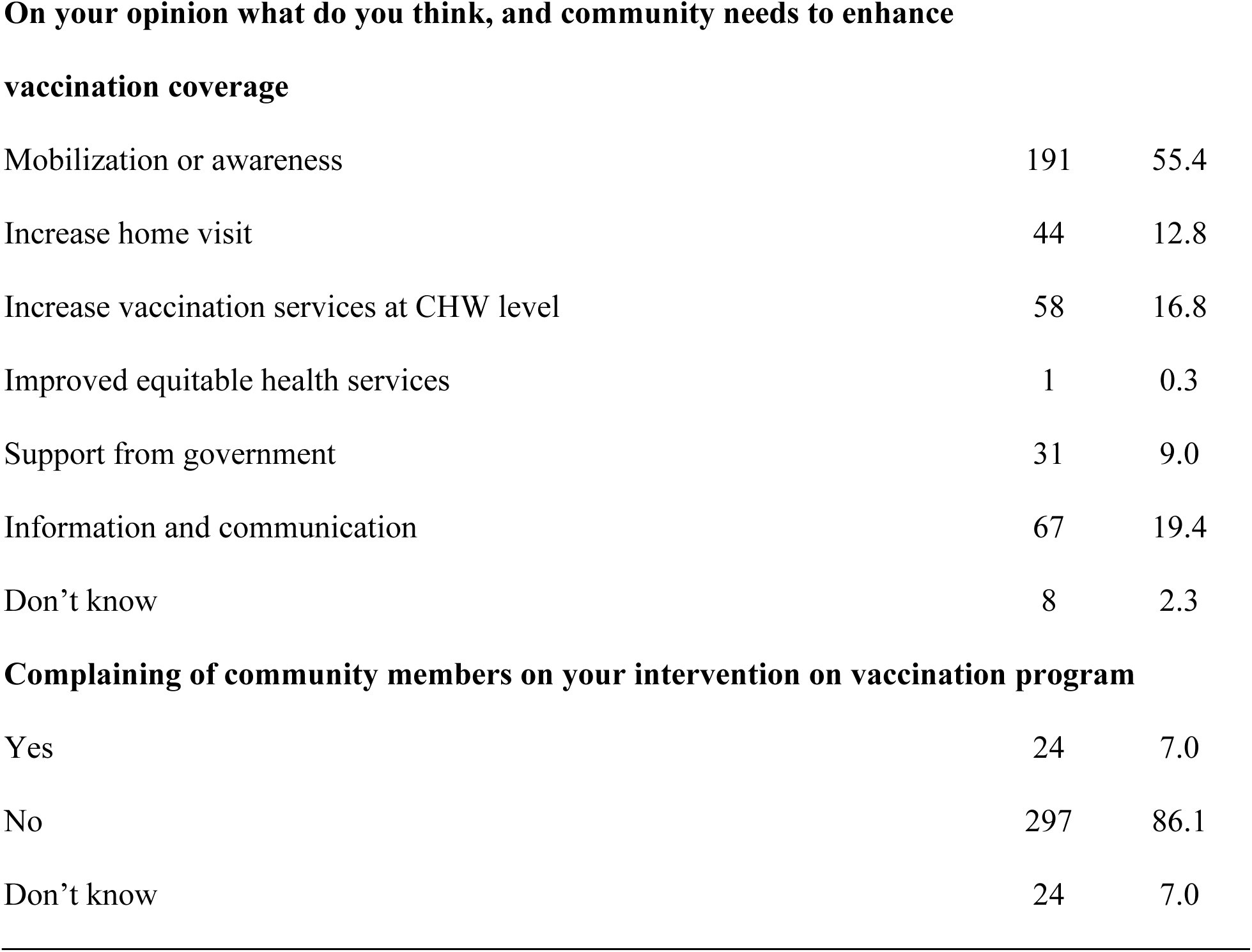
Community mobilization and participation towards childhood vaccination program.

Majority (61.4%) reported that community members were fully involved when vaccination event was planned. The main responsibilities of CHWs included referring children to healthcare facilities (58.0%) and administering vaccines during home visits (42.0%). Similarly, these findings are reflected in the qualitative data where the pregnant women interviewed indicated that they get advocacy and support through providing vaccinations and follow-ups as well as reminding them for vaccination appointment. Some of the pregnant women’s quotations were as follows:

> *“What is great is that they advocate in taking care of my child and reminding me to have my child vaccinated according to the given appointment* (KII 1)”.
>
> “*A community health worker visits our home, informs us about new vaccines, and advises us to take our child for vaccination* (KII-4)”; and another woman indicated “*It makes me happy when they bring vaccinations promptly or provide necessary information directly to my home* (KII-5)”.

Over half (55.4%) of the CHWs believed that mobilization or awareness to be conducted in order to enhance vaccination coverage. About 7.0% of the CHWs indicated that community members complained about vaccination. It was also reported that there are still ignorance and poor understanding according to the pregnant women participated in the qualitative study. These expressions include:

> “*It is due to poor understanding and lack of education. Families who are better informed understand the importance of vaccinations and ensure their children receive them*”. KII-4
>
> “*Some parents do not understand how vital vaccinations are because they lack proper education on the topic. Those who are educated take it seriously and follow through with the schedule*.” KII-1
>
> “*I cannot understand why a child wouldn’t be vaccinated if they are present. Perhaps some parents refuse vaccinations, but I cannot comprehend their reasoning*”. KII-5

### Infrastructure and logistics on childhood vaccination program

The primary methods of communication and transportation for follow-ups on child vaccination appointments were physical visits (89.6%), predominantly conducted on foot (99.7%). All CHWs reported covering their own communication expenses. All vaccines were stored and distributed exclusively from health centers (Table 4).

**Table 4.** Infrastructure and logistics on childhood vaccination program.

| Variables | N = 345 | % |
| --- | --- | --- |
| **Communication with caregivers during follow up on child vaccination appointment** |  |  |
| Call on phone | 5 | 1.4 |
| Physical visit | 309 | 89.6 |
| Others* | 31 | 9.0 |
| **Means of transport during home visit** |  |  |
| Bicycle | 1 | 0.3 |
| Foot | 344 | 99.7 |
| **Cover communication expenses/money** |  |  |
| Own | 345 | 100.0 |
| **Vaccines storage before distributed** |  |  |
| Health center | 345 | 100.0 |
| *Others: General village meeting, community services (umuganda), and during saving group meeting |  |  |

However, limited vaccination sites and long distance emerged as the key infrastructure and logistics barriers on implementing of childhood vaccination program. They indicated that in a cell there is only one vaccination site, which is limited vaccination site. The distance is identified as a burden for those coming from afar and is not easy for them to travel long distances just for a vaccination. This is how the key informants expressed their views:

> “*The problem is that there is just one vaccination site per cell. It makes it hard for everyone to reach it, especially those who live far away* (KII-3).”
>
> “*For families living far away, getting to the vaccination site is a real struggle. The distance makes it difficult for them to attend appointments on time* (KII-1).
>
> ” To overcome these challenges the pregnant women suggested that all the vaccines should be administered by CHWs “*Maybe some CHWs should be in charge and administrate the vaccines so that the issue get resolved and people with those issues be included*.” KII-3

### Suggestions achieving remaining 5% vaccination rate

When asked about recommendations for achieving full vaccination rate, CHWs prioritized monetary incentives or monthly salary along with additional trainings (51.9%) as the top suggestions. This was followed by increased mobilization about the importance of vaccination programs (13.3%) and provision of some materials such as digital tools (7.2%) as shown in Figure 1. These findings are further reinforced by the qualitative data from pregnant women, as demonstrated by the following quotes:

> *“I think CHWs are always complaining that they do not get any form of incentives or support when they visit pregnant women. It discourages them from doing their work effectively.” KII-5*
>
> *“A message should be sent as a reminder, not like in the past when it was just papers with dates. With technology, a mobile text message would be much better. It would help a mother to remember, instead of relying on papers.” KII-4*
>
> *We need more mobilization to help people understand how important vaccinations are for our children. KII-3*

**Figure 1.**
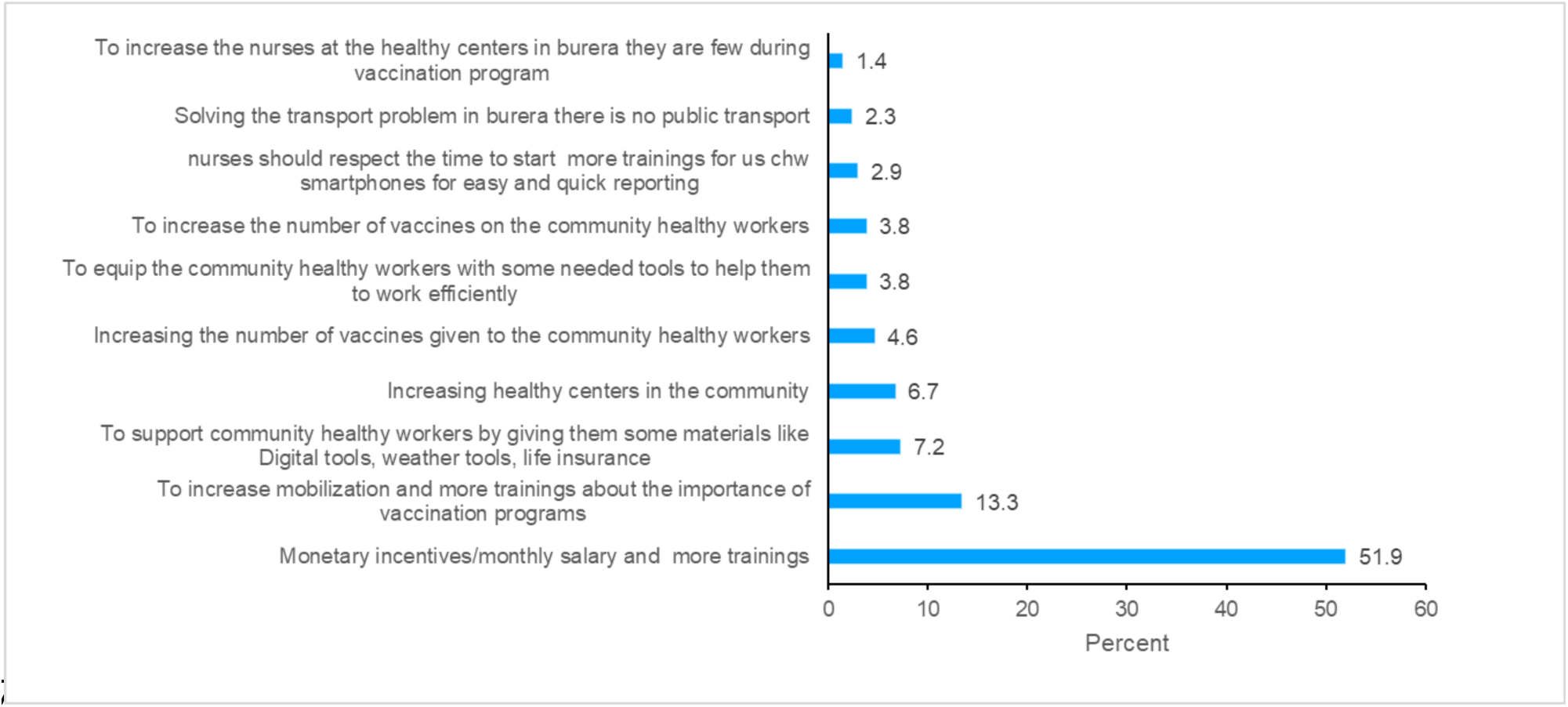
Suggests achieving remaining 5% vaccination rate.

## DISCUSSION

Given the persistent challenges of equitable access, vaccine hesitancy and health worker shortages, leveraging CHWs as vaccinators is an important opportunity ^16^. The deployment of Community Health Workers (CHWs) is widely promoted as a strategy for reducing health inequities in low- and middle-income countries (LMIC) ^17^.

The study found that the majority of CHWs were female (67.5%) and aged between 40 and 49 years (38.8%), with primary education being the highest level attained by most (86.1%). These demographic characteristics align with previous studies in sub-Saharan Africa, where CHWs are predominantly middle-aged women with limited formal education but extensive community engagement ^18^. The dominance of women among CHWs is critical, as they are more likely to communicate effectively with mothers and caregivers about childhood vaccination. However, the limited educational background of CHWs may affect their ability to effectively deliver health messages, a challenge that has been noted in other studies ^19^. Our study highlights that 94.5% of CHWs received training on vaccination services, with most (64.1%) attending training at least once or twice a year, and this supporting the argument of WHO that adequate training is essential for effective immunization programs. These findings are consistent with studies in in Uganda and Kenya, which emphasize the importance of continuous CHW training to improve knowledge and vaccination uptake ^20^. Qualitative responses from caregivers also emphasized the effectiveness of CHWs in disseminating vaccination information, which echoes findings from a study that highlighted CHWs’ role in bridging knowledge gaps among mothers ^21^.

The findings indicated that CHWs significantly contribute to increasing immunization rates through education, household visits, and follow-up which align with previous research that highlights the crucial role of CHWs in increasing childhood vaccination coverage. Similar to a study conducted in Ethiopia, which found that CHWs played a pivotal role in immunization services, our results confirm that CHWs actively engage in community mobilization and follow-up visits ^22^. The predominance of female CHWs in our study (67.5%) also mirrors global trends, where women often take the lead in community health interventions due to their trusted roles within communities ^17^. The qualitative findings further reinforce the effectiveness of CHWs in ensuring timely vaccination, as echoed by caregivers who acknowledged the proactive role of CHWs in reminding them about vaccination schedules. This aligns with research from Nigeria, where CHWs’ home visits significantly increased vaccine uptake ^20^.

The study found that 93.9% of CHWs were fully involved in vaccination campaigns, a crucial factor in vaccine acceptance and coverage. This is in line with findings from Ghana, where strong community engagement through CHWs improved childhood immunization rates. Additionally, over half (55.4%) of CHWs emphasized the need for continued mobilization efforts to enhance vaccination coverage. Similar findings were reported in Kenya, where CHWs’ proactive community engagement led to increased routine immunization uptake ^22^. However, 7.0% of CHWs reported resistance from community members, often due to misinformation and lack of awareness, which is a common barrier in many low-resource settings ^19^.

Despite their critical role, CHWs faced logistical challenges, with 99.7% relying on foot transport and covering communication expenses themselves. Vaccine storage was centralized at health centers, and only one vaccination site existed per cell, making access difficult for remote populations. Studies in Uganda and Nigeria have similarly highlighted transportation and cold chain constraints as key barriers to effective vaccine delivery ^23^. The limited vaccination sites, as indicated by the qualitative data, reinforce the need for decentralizing vaccination services to improve accessibility ^24^. The perspectives of mothers and caregivers in this study further reinforce the importance of CHWs in vaccination programs. The qualitative findings revealed that CHWs are trusted sources of information and play a key role in appointment reminders, follow-ups, and vaccine advocacy. These findings are consistent with previous studies highlighting the trust placed in CHWs as frontline healthcare providers in rural settings ^25^. However, caregivers also identified gaps in community understanding, with some parents still showing reluctance due to misinformation or lack of education. Similar hesitancy has been reported in other studies, emphasizing the need for intensified awareness campaigns ^26^.

### Challenges and recommendations for improving vaccination coverage

Infrastructure and logistical barriers emerged as major constraints, particularly the limited number of vaccination sites and the long distances some families must travel. This is consistent with findings from other studies where inadequate access to vaccination services, community awareness and misinformation was identified as a leading factor in incomplete immunization ^26^. Additionally, CHWs reported a lack of incentives, which negatively affects their motivation. Studies have shown that providing CHWs with financial incentives or non-monetary support, such as mobile technology for reminders, significantly enhances their effectiveness ^27^. Further, the frequency of training remains relatively low, with 64.1% of CHWs receiving training not more than twice in year, suggesting the need for enhanced refresher courses to maintain competency. To address these challenges, CHWs recommended increased awareness campaigns, government support, and financial incentives. These recommendations align with global best practices, where strengthening CHW training, providing stipends, and integrating digital health tools have proven to enhance vaccination outcomes ^28^. Mobile technology, such as SMS reminders for vaccination appointments, was also suggested and has been successfully implemented in other low-resource settings to improve compliance rates ^29^.

## CONCLUSION

The findings of this study highlight the significant role of CHWs in promoting childhood vaccination and the barriers they face. While CHWs have positively influenced vaccination coverage through mobilization and home visits, infrastructural and logistical challenges remain. Strengthening CHW training, increasing incentives, and improving access to vaccination services could enhance childhood immunization rates. Future research should explore the long-term impact of CHW-led interventions and innovative solutions such as digital health tools to optimize vaccination efforts in rural settings. Findings provide insights for policymakers and communities, helping to bridge the last mile of remaining 5% vaccination coverage by 2030.

## Ethical approval

This study was approved by the University of Global Health Equity (UGHE) Institutional Review Board on the 9th of April 2024 (reference number UGHE-IRB/2024/240). Prior to participation, all participants provided written informed consent.

## Data Availability

All data contained within the main body of the text are publicly available.

